# Impact of ABO blood groups on residual pulmonary vascular obstruction in patients with unprovoked pulmonary embolism

**DOI:** 10.1101/2025.10.09.25337585

**Authors:** Floriane Samaria, Ohanna C. L. Bezerra, Lénaïck Gourhant, Marine Germain, Gaëlle Munsch, Robert Olaso, Delphine Bacq, Catherine A Lemarié, Grégoire Le Gal, Jean-François Deleuze, France Gagnon, Marc A Rodger, Francis Couturaud, David-Alexandre Trégouët

## Abstract

**Background:** After completing anticoagulant therapy for acute pulmonary embolism (PE), patients may have persistent thrombotic materials in the lung, causing residual pulmonary vascular obstruction (RPVO). Detectable by imaging, RPVO is associated with an increased risk of recurrent events and chronic complications. Identifying RPVO risk factors has become a recent focus, yet no genetic determinants have been established. This study specifically investigates the association between genetically determined ABO blood groups and RPVO in patients with unprovoked-PE.

**Methods:** This work relies on two French cohorts of patients with unprovoked-PE, EDITH and PADIS-PE, and a Canadian one, REVERSE-I, where RPVO was measured with the same protocol. In each study, five *ABO* polymorphisms were used to infer *ABO* blood groups: rs8176719-delG, rs41302905-T, rs2519093-T, rs1053878-A and rs8176743-T alleles tagging for O1, O2, A1, A2 and B, respectively. Associations between ABO blood groups and RPVO were assessed in each study using a Compound Poisson Gamma model, which accounts for the semi-continuous RPVO distribution. Study-specific results were then combined through meta-analysis.

**Results:** The meta-analysis included 586 patients with unprovoked-PE, providing sufficient power (>80%), as confirmed by simulations, to detect moderate genetic effects (1.3-fold increased risk) of common variants (allele frequency ≥0.05). Despite this, no significant association was observed between ABO blood groups and RPVO, and no consistent effect was seen across the three studies.

**Interpretation:** ABO blood groups are unlikely to be major risk factors for RPVO. Nevertheless, this study provides an important foundation for future large-scale genetic investigations aimed at identifying the molecular determinants of RPVO.

## Introduction

Pulmonary embolism (PE) is the third most frequent cause of cardiovascular death in the world, right after myocardial infarction and stroke (1–3). As the most severe clinical manifestation of venous thromboembolism (VTE), PE is associated with a high short-term mortality rate, with nearly one in four patients experiencing sudden death in high-risk groups of patients (4). Fortunately, the short-term prognosis of PE has markedly improved in recent years driven by advances in diagnosis, risk-stratification and treatments (5), leading to lower mortality rates of PE patients over the years (1). Although the management of PE primarily focuses on its acute manifestation, the development of chronic long-term complications raises new challenges in PE management (6). This is especially evident in unprovoked-PE, defined as PE occurring in the absence of any major identified risk factors (7,8). Indeed, after 3 to 6 months of standard anticoagulant therapy, about 30% of unprovoked-PE patients still have residual thrombotic material that did not fully resolve in the pulmonary arteries, forming a persistent obstructive scars adhered to their walls (9,10). Unless surgically removed, this residual pulmonary vascular obstruction (RPVO) persists within the pulmonary circulation. RPVO measured after 6 months of uninterrupted anticoagulation therapy has been shown to be a strong predictor of VTE recurrence in patients with a first unprovoked-PE (10–12). Moreover, in 10% of unprovoked-PE cases, RPVO is associated with the development of chronic thromboembolic pulmonary hypertension (CTEPH) (13). These patients have a reduced life expectancy, with an estimated 3-year survival of ∼90% for those undergoing surgery (i.e., thromboendarterectomy, the only curative treatment for CTEPH) and 70% for those who are deemed inoperable (13).

The pathophysiology of RPVO remains poorly understood. Despite growing interest, the underlying mechanisms causing RPVO, recurrent VTE, or CTEPH are still unclear. Recent research has increasingly focused on identifying RPVO risk factors and elucidating its underlying mechanisms. To date, advanced age, elevated initial obstruction percentage at PE diagnosis and some fibrinogen properties have been reported as risk factors and remain the only known predictors (5,6,14–17). *ABO* is a major genetic risk factor for first VTE (18), with a stronger association reported for deep vein thrombosis (DVT) than for PE (19). *ABO* is also linked to VTE recurrence risk (20). In DVT patients, non-O blood groups have been associated with residual vein obstruction (21). Furthermore, CTEPH patients more frequently carry non-O blood groups than those with non-thromboembolic pulmonary hypertension (22). However, the association of ABO blood groups with RPVO in PE patients has never been explored. This study addresses this gap by examining the relationship between genetically determined ABO blood groups and RPVO across three independent clinical studies.

## Methods

### Studies and participants

This work builds upon two French studies, EDITH (23–26) and PADIS-PE (11,27), as well as one Canadian study, REVERSE-I (28–30).

EDITH (Etude des Déterminants/Interaction de la Thrombose veineuse) is a prospective observational cohort study launched in 2000 aimed at improving the understanding of VTE risk factors. Since its inception, it has enrolled more than 5,000 VTE patients. For the current work, we focused on patients over 18 years of age who experienced a first episode of unprovoked-PE, completed 3 to 6 months of anticoagulation therapy, underwent a ventilation/perfusion (V/Q) scan within 3 to 24 months after treatment, and had available DNA materials. PE was defined as unprovoked in absence of the following factors in the past 3-months or past years before PE: cancer, pregnancy, postpartum, oral contraceptive use, immobilization on the inferior member, and surgery involving long immobilization during or after operative phase. The EDITH study was approved by Brest University Hospital scientific and ethics board, in accordance with the Declaration of Helsinki. Written consent for the study and DNA analysis was obtained from all patients.

PADIS-PE (Prolonged Anticoagulant During Eighteen Months vs Placebo After Initial Six-month Treatment for a First Episode of Idiopathic Pulmonary Embolism) is a randomized clinical-trial (NCT00740883) conducted between 2007 and 2014 in 14 French care centers. PADIS-PE enrolled 371 unprovoked-PE cases during this period. The goal was to identify the benefits and drawbacks of 18 months of treatment versus 6 months. Patients over 18 years old, with a first episode of unprovoked-PE and V/Q scan available right after 6 months of uninterrupted treatment were included. All exclusion criteria are set out in the study protocol (11). PADIS-PE study was conducted in accordance with the ethical principles stated in the Declaration of Helsinki, Good Clinical Practice, and relevant French regulations regarding ethics and data protection. The protocol and amendments were approved by a central independent ethics committee and written informed consent was obtained from all participants.

REVERSE-I (Recurrent Venous Thromboembolism Risk Stratification Evaluation) is a Canadian multicentric prospective study which enrolled 646 patients between 2001 and 2006 after their first episode of VTE. The aim was to derive a new clinical decision rule that would facilitate identification of patients at low risk of recurrent VTE after completion of treatment. All exclusion criteria can be found elsewhere (29,30). Patients were included in the current study if they were aged over 18, with V/Q scan available right after completion of their 5-7 months oral anticoagulant therapy and had a first unprovoked-PE event. Unprovoked-PE was defined as PE occurring in the absence of the following: leg fracture or lower extremity plaster cast, immobilization for more than three days or surgery using a general anesthetic in the three months prior to the index event, diagnosis of malignancy in the past five years at the time of enrollment, pregnancy or six weeks postpartum statement, use of oral contraceptive and hormone replacement. Institutional research ethics board approvals were obtained by all participating centers (Ottawa Hospital Research Ethics Board). Protocols to study genetics in REVERSE I was approved by the University of Toronto Research Ethics Board.

In total, 165 unprovoked-PE patients from the EDITH study were included, all of whom had available data on both RPVO and genetically determined ABO blood groups. Corresponding numbers were 306 and 115 in PADIS-PE and REVERSE-I, respectively.

### RPVO measurements

In the three contributing studies, V/Q scans were performed to measure RPVO values. The protocol proposed by Meyer et al (31) to correctly assess RPVO from these measures was adopted. Independent investigators scored mismatched perfusion defects by examining air and blood flow in the lungs (5,32) to obtain a global score of obstruction (in percentage). Briefly, radioactive tracers are intravenously injected and blocked on their first passage through the pulmonary capillaries so that imaging is readable. A perfusion score is then attributed to each five lung’s lobes, to whom a weight based on the percentage of vascularization is primarily given. Then a global perfusion score is obtained by summing all the perfusion scores individually multiplied by their respective weights. Finally, the percentage of RPVO is obtained by subtracting to 1 the global perfusion score (S) such that RPVO = (1-S) × 100. A threshold of ≥5% is commonly used to define the presence of RPVO. However, its value varies across studies (12,26,33–37), and most investigations on RPVO have been case–control studies employing different obstruction thresholds. To date, no consensus has been reached within the scientific and medical community regarding the optimal cutoff, although Picart et al. showed that a ≥5% threshold is sufficient to characterize RPVO (23). In the present study, we use a continuous measurement of RPVO, allowing us to account for the degree of obstruction in each patient, since RPVO values can range from 0 to 100.

### ABO blood groups determination

ABO blood groups were genetically determined from 5 common ABO single nucleotide polymorphisms (SNPs) as described in Goumidi et al (18): rs8176719-delG (tagging O1), rs41302905-T (O2), rs2519093-T (A1), rs1053878-A (A2), and rs8176743-T (B).

As DNA samples from EDITH, PADIS-PE and REVERSE-I participants were typed for high-density genotyping arrays, we used best-guessed genotypes from imputed data to infer ABO blood groups. The description of genotyping, imputation and derivation of principal components from genome wide genotype data in each of the three studies have already been provided in Munsch et al (38). Briefly, individuals from EDITH and PADIS-PE were genotyped by the Illumina Infinium Human Global Screening Array GSAMD-24v3 at the French National Center of Human Genomics Research and data were further imputed on the 1000 genomes phase 3v5 panel. REVERSE-I participants were genotyped on the Illumina Global Screening Array at Génome Québec and genotype data further imputed using TOPmed Eagle v2.4 panel. The imputation quality scores of ABO blood groups tagging SNPs in each of the three studies are shown in **Supplementary Table 1**. All five ABO tagging SNPs have imputation quality greater than 0.90 across all three studies, except the uncommon O2 tagging variant with imputation quality of 0.83 in REVERSE-I.

### Statistical Analyses

For descriptive analyses, continuous variables were expressed as mean and standard deviation. Categorical variables were expressed as numbers and percentages.

In each of the three contributing studies, the distribution of RPVO measurements was characterized by an excess of zero values followed by positive continuous values (**Figure 1 and Table 1**). Consequently, we used a Compound Poisson-Gamma (CPG) model (39,40) to investigate the association of clinical and biological variables (including age, sex, body mass index (BMI), PE associated or not with DVT and ABO blood groups) with RPVO. In a CPG modeling, the exponential of a regression coefficient represents the multiplicative change in the RPVO measurement associated with one-unit increase in the given covariate. Association analyses were conducted in each study while adjusting for age and sex. Results were then meta-analyzed across studies using the inverse-variance method (41,42).

**Figure 1.**
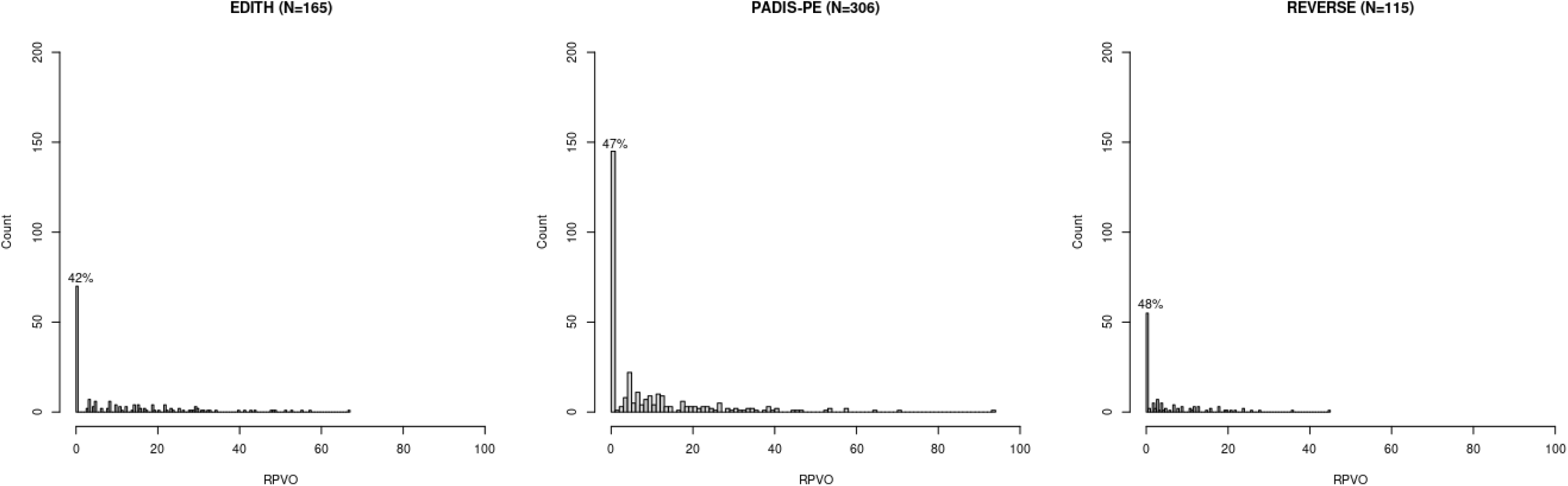
Distributions of RPVO and respective percentages of zero values in unprovoked-PE patients from EDITH, PADIS-PE, and REVERSE-I.

**Table 1.** Distribution of zeros and positive values of RPVO in unprovoked-PE patients from EDITH, PADIS-PE and REVERSE-I.

|  | **EDITH**<br>N = 165 | **PADIS-PE**<br>N = 306 | **REVERSE-I**<br>N=115 | **Combined studies**<br>N=586 |
| --- | --- | --- | --- | --- |
| **RPVO=0** (N, %) | 70 (42.4) | 144 (47.0) | 55 (47.8) | 269 (45.9) |
| **RPVO>0** (N, %) | 95 (57.6) | 162 (53.0) | 60 (52.2) | 317 (54.1) |
| **Mean** (sd)* | 19.6 (14.6) | 17.1 (15.3) | 10.4 (9.1) | 15.5 (14.4) |
| **Q1-Q2-Q3\*** | 8.2-15.5-28.2 | 6.2-11.4-23.2 | 3.0-7.5-15.2 | 6.0-11.7-22.5 |

\*First, second and third quartiles for positive RPVO values.

The association analysis of ABO blood groups with RPVO was conducted assuming additive allele effects and was adjusted for the first four principal components derived from the genotypic data. The O1 blood group was used as reference. Statistical analyses were performed using *R* (v4.1.0), using the *cplm* (v0.7.12) package for CPG analyses and using *metafor* (v4.6-0) package for fixed effect meta-analysis.

## Results

The main subject characteristics are shown in **Table 2**. A total of 165 unprovoked-PE with RPVO measurements and genetic data available were reported in EDITH, 306 in PADIS-PE and 115 in REVERSE. Some common characteristics were observed across the three studies, with comparable proportions of women and mean body mass index (BMI). Approximately 50% of participants are women in the three studies, the mean BMI was around 28 kg/m². The distribution of RPVO did not significantly differ (p=0.12) across the three studies, despite some trend for quartiles of RPVO being lower in REVERSE-I. On the other hand, some differences were detected, reflecting distinctions between EDITH and the more similar PADIS-PE and REVERSE-I. Patients from EDITH are older on average (65.8 years old) than patients from PADIS-PE (59.3 years old) and REVERSE-I (54.6 years old). Similarly, the proportion of DVT associated with unprovoked-PE was higher in EDITH (48.5%) compared to PADIS-PE (30.8%) and REVERSE-I (27.5%). The same pattern was observed for the proportion of VTE recurrent and death events. Regarding the time to recurrence and follow up, EDITH and REVERSE-I displayed longer times to events (2.6 years and 5.4 years for recurrence; 6.6 years and 5.9 years for death events, respectively) than PADIS-PE (1.0 years and 1.58 years).

**Table 2.** Characteristics of the unprovoked-PE patients from EDITH, PADIS-PE and REVERSE-I.

|  | **EDITH**<br>N = 165 | **PADIS-PE**<br>N = 306 | **REVERSE-I**<br>N=115 |
| --- | --- | --- | --- |
| **Age** (years, mean (sd)) | 65.8 (14.3) | 59.3 (17.3) | 54.6 (17.7) |
| **Female** (N,%) | 89 (52.1) | 158 (51.6) | 54 (46.9) |
| **BMI** (kg/m<sup>2</sup>, mean (sd)) | 27.7 (6.1)<br>NA=4 | 27.6 (5.3) | 29.3 (6.9)<br>NA=1 |
| **DVT associated** (yes, (N,%)) | 80 (48.5) | 92 (30.8)<br>NA=8 | 30 (27.5)<br>NA=6 |
| **CTEPH\*** (yes, (N,%)) | 4 (2.6)<br>NA=11 | 8 (2.6) | - |
| **Recurrent event of VTE** (yes, (N,%)) | 39 (26.0)<br>NA=15 | 57 (18.6)<br>NA=35 | 19 (16.5) |
| **Time to recurrence** (years, mean (sd)) | 2.6 (2.9) | 1.8 (1.0) | 5.4 (3.7) |
| **Death events** (N (%)) | 33 (20.1)<br>NA=1 | 14 (5.3)<br>NA=40 | 8 (6.9) |
| **Time of follow up** (years, mean (sd)) | 6.6 (2.9) | 1.58 (1.1) | 5.9 (2.8) |
| **RPVO** (Q1-Q2-Q3) | 0 - 5.0 - 18.7 | 0 - 3.2 - 12.2 | 0 - 2.0 - 8.0 |

\*Chronic thromboembolic pulmonary hypertension (CTEPH) was assessed in EDITH for patients with a high-probability V/Q lung scan, a mean pulmonary artery pressure >25mmHg at rest, and a pulmonary wedge pressure <15mmHg on right heart catheterization. In PADIS-PE, the same criteria were used, with the addition of RPVO >5%. The information was not available in REVERSE-I. NA = Not available.

Results of the association of clinical/biological factors with RPVO are shown in **Table 3**. Overall, age was the only variable found to be significantly associated with RPVO with homogenous effect across the three studies. Specifically, each additional year of age was associated with a 4% increase in RPVO (exp(β)=1.04, p=5.56×10^−7^ in the meta-analyzed samples). No other variables were associated with RPVO measurements.

**Table 3.**
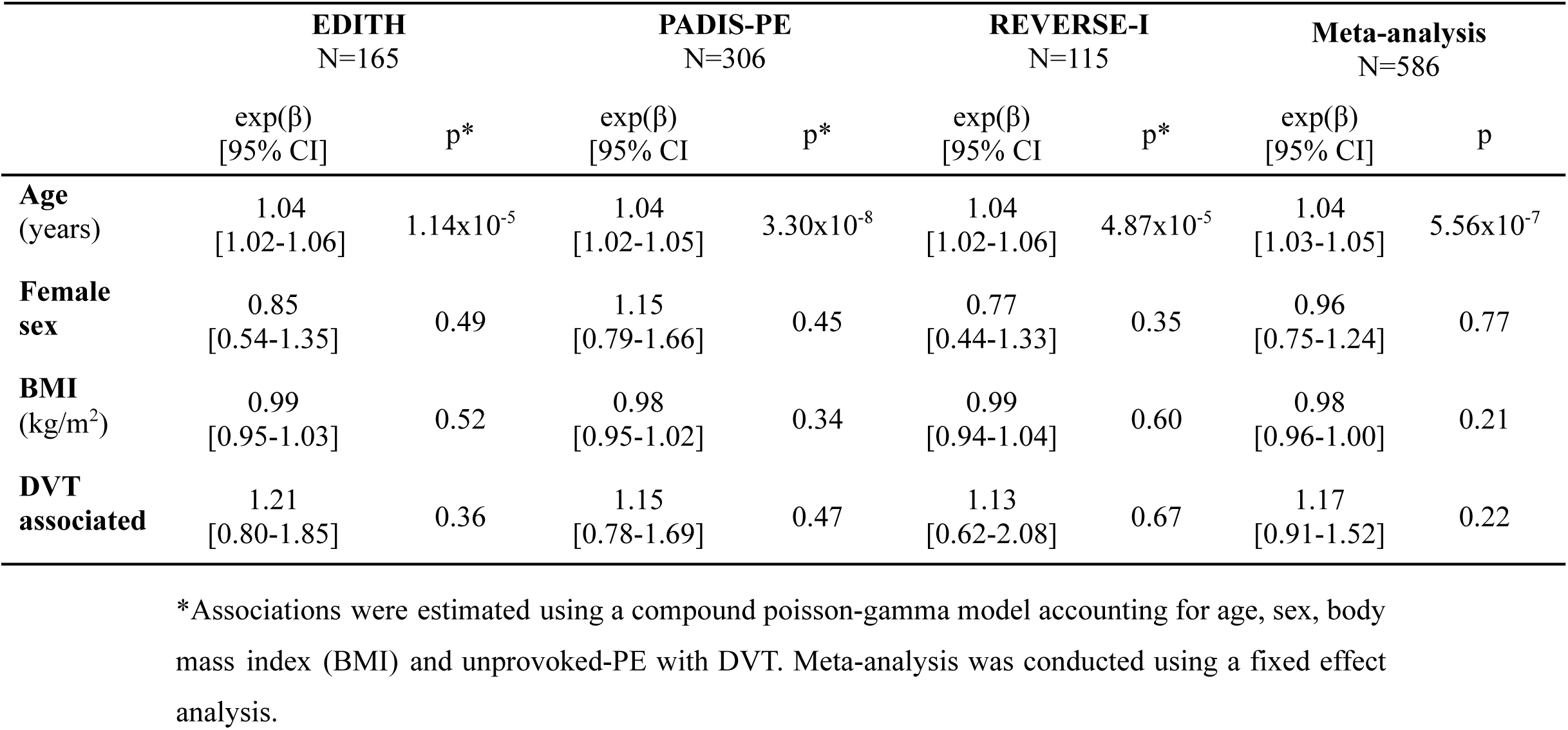
Clinical and biological correlates of RPVO in unprovoked-PE patients from EDITH, PADIS-PE and REVERSE-I.

|  | **EDITH**<br>N=165 |  | **PADIS-PE**<br>N=306 |  | **REVERSE-I**<br>N=115 |  | **Meta-analysis**<br>N=586 |  |
| --- | --- | --- | --- | --- | --- | --- | --- | --- |
| | exp($\beta$)<br>[95% CI] | p* | exp($\beta$)<br>[95% CI] | p* | exp($\beta$)<br>[95% CI] | p* | exp($\beta$)<br>[95% CI] | p |
| **Age**<br>(years) | 1.04<br>[1.02-1.06] | 1.14x10<sup>-5</sup> | 1.04<br>[1.02-1.05] | 3.30x10<sup>-8</sup> | 1.04<br>[1.02-1.06] | 4.87x10<sup>-5</sup> | 1.04<br>[1.03-1.05] | 5.56x10<sup>-7</sup> |
| **Female sex** | 0.85<br>[0.54-1.35] | 0.49 | 1.15<br>[0.79-1.66] | 0.45 | 0.77<br>[0.44-1.33] | 0.35 | 0.96<br>[0.75-1.24] | 0.77 |
| **BMI**<br>(kg/m<sup>2</sup>) | 0.99<br>[0.95-1.03] | 0.52 | 0.98<br>[0.95-1.02] | 0.34 | 0.99<br>[0.94-1.04] | 0.60 | 0.98<br>[0.96-1.00] | 0.21 |
| **DVT associated** | 1.21<br>[0.80-1.85] | 0.36 | 1.15<br>[0.78-1.69] | 0.47 | 1.13<br>[0.62-2.08] | 0.67 | 1.17<br>[0.91-1.52] | 0.22 |

\*Associations were estimated using a compound poisson-gamma model accounting for age, sex, body mass index (BMI) and unprovoked-PE with DVT. Meta-analysis was conducted using a fixed effect analysis.

Results of the association analysis of ABO haplotypes with RPVO are displayed in **Figure 2**, **Table 4 and Table 5**. **Table 4** presents the estimated ABO haplotypes frequency distribution in patients with RPVO=0 and in those with positive (non-zero) RPVO. Haplotype frequencies were similar across the three studies and did not differ between patients with or without RPVO, as shown in **Supplementary Table 2**. **Table 5** provides detailed association results from applying the CPG model across the three studies while **Figure 2** displays a forest plot summarizing the overall findings. In EDITH, a marginal (p=0.01) association was observed between the A2 blood group and RPVO compared with O1, but this trend was not found in the other studies. No significant associations were seen for the other ABO haplotypes, and the combined meta-analysis indicated that ABO haplotypes have no significant impact on RPVO. We also performed similar association analyses of ABO haplotypes with RPVO, defined according to commonly used binary thresholds in previous studies. Regardless of the threshold applied, no association was detected. (**Supplementary Table 3**).

**Figure 2.**
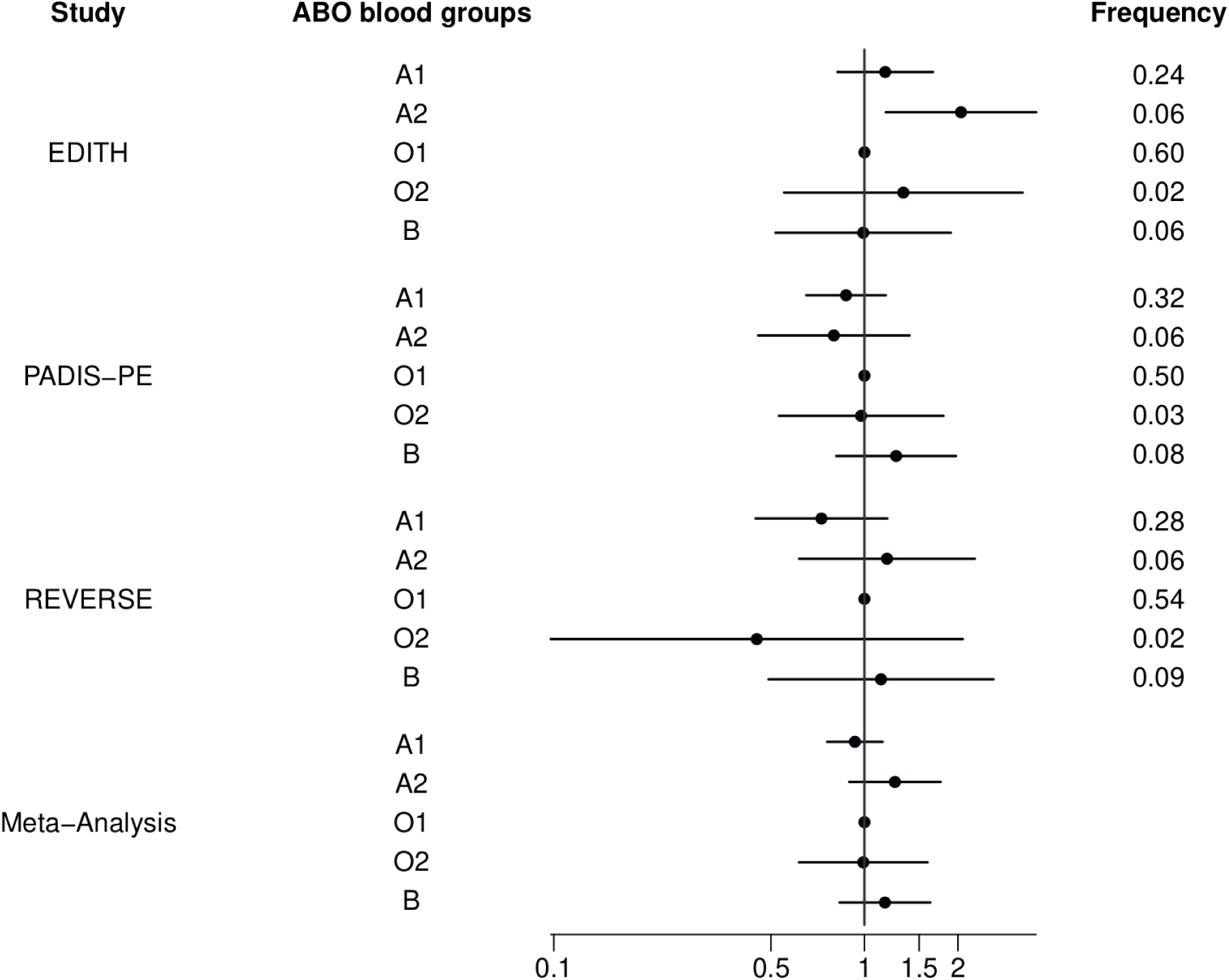
Forest plot of associations of ABO blood groups with RPVO in unprovoked-PE patients from EDITH, PADIS-PE, and REVERSE-I. Associations were tested using a compound poisson-gamma model adjusted on age, sex and the four first principal components, under the assumption of additive allele effect. O1, taken as the reference group, was tagged by the rs8176719-delG allele. The O2, A1, A2 and B blood groups were tagged by the rs41302905-T, rs2519093-T, rs1053878-A and rs8176743-T alleles, respectively.

**Table 4.** ABO haplotype frequencies (%) by RPVO status in unprovoked-PE patients from EDITH, PADIS-PE and REVERSE-I.

| ABO<br>haplotypes | **EDITH**<br>N=165 |  | **PADIS-PE**<br>N=306 |  | **REVERSE**<br>N=115 |  | **Combined studies**<br>N=586 |  |
| --- | --- | --- | --- | --- | --- | --- | --- | --- |
|  | RPVO=0 | RPVO>0 | RPVO=0 | RPVO>0 | RPVO=0 | RPVO>0 | RPVO=0 | RPVO>0 |
| O1 | 62.32 | 59.47 | 46.86 | 53.13 | 54.54 | 54.24 | 52.44 | 55.26 |
| O2 | 2.17 | 3.16 | 2.10 | 4.06 | 3.64 | 1.69 | 2.43 | 3.34 |
| A1 | 20.29 | 27.37 | 37.06 | 29.38 | 29.09 | 26.27 | 31.09 | 28.19 |
| A2 | 7.25 | 4.74 | 7.34 | 4.37 | 3.64 | 9.32 | 6.55 | 5.41 |
| B | 7.97 | 5.26 | 6.64 | 9.06 | 9.09 | 8.48 | 7.49 | 7.80 |

**Table 5.** Associations of ABO haplotypes with RPVO in unprovoked-PE patients from EDITH, PADIS-PE, and REVERSE-I.

| ABO<br>haplotypes | EDITH<br>N=165 |  | PADIS-PE<br>N=306 |  | REVERSE<br>N=115 |  | Meta-analysis<br>N=586 |  |
| --- | --- | --- | --- | --- | --- | --- | --- | --- |
| | exp($\beta$) [95%<br>CI] | p* | exp($\beta$) [95% CI] | p* | exp($\beta$) [95%<br>CI] | p* | exp($\beta$) [95%<br>CI] | p |
| O1 | Reference |  | Reference |  | Reference |  | Reference |  |
| O2 | 1.33 [0.55-3.23] | 0.52 | 0.97 [0.53-1.79] | 0.94 | 0.45 [0.09-2.07] | 0.30 | 0.99 [0.61-1.59] | 0.97 |
| A1 | 1.16 [0.82-1.66] | 0.39 | 0.87 [0.65-1.17] | 0.36 | 0.73 [0.45-1.18] | 0.20 | 0.93 [0.76-1.14] | 0.50 |
| A2 | 2.04 [1.17-3.57] | 0.01 | 0.79[0.46-1.40] | 0.43 | 1.18 [0.62-2.26] | 0.62 | 1.25 [0.89-1.75] | 0.19 |
| B | 0.99 [0.52-1.89] | 0.97 | 1.26 [0.81-1.97] | 0.30 | 1.13 [0.49-2.60] | 0.77 | 1.16 [0.83-1.63] | 0.37 |

\*Associations were tested using a compound poisson-gamma model adjusted on age, sex and the four first principal components, under the assumption of additive allele effect.
O1, taken as the reference group, was tagged by the rs8176719-delG allele. The O2, A1, A2 and B blood groups were tagged by the rs41302905-T, rs2519093-T, rs1053878-A and rs8176743-T alleles, respectively.

## Discussion

To our knowledge, this is the first study to assess the role of genetically determined ABO blood groups with RPVO in patients with unprovoked-PE. Moreover, it is the first to apply the CPG model to analyze RPVO measurements while accounting for the semi-continuous nature of this clinical phenotype. This statistical approach has recently been shown to be well suited for genetic association studies of continuous variables with excess of zero values (40).

While Dentali et al. reported that non-O blood groups were associated with a three-fold increased risk of RVO in 268 patients with DVT (21), we did not find any significant association of ABO blood groups with RPVO in unprovoked-PE patients. With a sample size of 586 patients, we had approximately 90% statistical power (α=0.05) to detect a risk increase of at least 1.20 for non-O blood groups, assuming a frequency of 0.58 (**Supplementary Table 4 – Supplementary Methods**). For a single nucleotide polymorphism with a MAF as low as 0.03—quite similar to the rare A2, B, and O2 blood group tagging variants—our study had approximately 80% power to detect an allelic effect corresponding to at least an 80% increase in risk. Similarly, our study had sufficient power (>80%) to detect an increased risk in RPVO greater than 1.30 for any polymorphism with MAF>0.20. These findings suggest that if ABO blood groups influence RPVO, the effect is likely more modest and would require much larger studies to be reliably detected. Bonderman et al. (22) reported that non-O blood groups were associated with a two-fold increased risk of CTEPH, for which RPVO is a known risk factor (6,15,43,44). The lack of association between ABO blood groups and RPVO observed in our study suggests that ABO may influence the risk of CTEPH via mechanisms independent of RPVO. We could not test this hypothesis in our study because the number of unprovoked-PE patients who developed CTEPH during the follow-up was too small (N=12). This hypothesis should be evaluated in a large clinical prospective cohort of PE patients with available DNA and RPVO measurements. Of note, exclusion of these two patients from the analysis did not alter the results of the haplotype association study (data not shown).

In conclusion, our study found no evidence of an association between ABO blood groups and RPVO in unprovoked-PE patients. Nonetheless, it lays important foundations for future large-scale genetic studies aimed at uncovering the molecular determinants of RPVO.

## Data Availability

All data produced in the present study are available upon reasonable request to the authors

## Acknowledgments

F.S is supported by the EUR DPH, a PhD program supported within the framework of the PIA3 (Investment for the future), project reference 17-EURE-0019. G.M. was partially supported by the EPIDEMIOM-VT Senior Chair from the University of Bordeaux initiative of excellence IdEX. G.L.G. holds a Distinguished Clinical Research Chair on Prevention and Diagnosis of Venous Thromboembolism from the Department of Medicine, University of Ottawa. Dr. Marc Rodger was supported by McGill University’s Harry Webster Thorpe Chair.

Genetic analyses in the REVERSE-I study were supported by the Canadian Institutes of Health Research (grant PJT-162233) and by a fellowship from the CANSSI Ontario STAGE (Genetic Epidemiology and Statistical Genetics Research training program). We are grateful to the Centre for Applied Genomics and The Hospital for Sick Children (Toronto, Canada) for their contributions to genotyping and data processing. Computational analyses of REVERSE I data were facilitated by the SciNET High Performance Computing Consortium at the University of Toronto and the Digital Research Alliance of Canada.

EDITH and PADIS-PE genomics research programs were supported by the GENMED Laboratory of Excellence on Medical Genomics [ANR-10-LABX-0013], a research program managed by the National Research Agency (ANR) as part of the French Investment for the Future. Statistical analyses of EDITH and PADIS-PE genomic data benefited from the technical support of the CBiB computing centre of the University of Bordeaux. In France, the EDITH and PADIS-PE studies are part of the French Clinical Research Infrastructure Network on Venous Thrombo-Embolism (F-CRIN INNOVTE). In Canada participating centers are members of the CanVECTOR Network. These national networks are members of INVENT-VTE, the International Network of Venous Thromboembolism Clinical Research Networks (www.invent-VTE.com).

This work was partially supported by the MORPHEUS (Prognosis iMprOvement of unpRovoked venous tHromboEmbolism Using perSonalized anticoagulant therapy) initiative, a multidisciplinary research project funded by the European Commission under the Horizon Europe programme (grant HORIZON-HLTH-2022-TOOL-11-01-2022).

## Authorship Contributions

M.R, F.C and D-A.T participated in study concept and design. C.L, L.G, M.R and F.C participated in phenotype data acquisition or control quality. O.B, F.G, R.O, A.B, D.B, J-F.D, G.M, M.G and D-AT participated in genotype data acquisition or control quality. F.S, O.B, G.M, F.G and D-A.T participated in data analysis and interpretation. F.S and D-AT wrote the initial draft of the manuscript which was reviewed and approved by all co-authors.

## Ethics approval

All experimental protocols to study the genetics of RPVO were approved by the local ethic committees:

- The EDITH study was approved by Brest University Hospital scientific and ethics board, in accordance with the Declaration of Helsinki.
- PADIS-PE study was conducted in accordance with the ethical principles stated in the Declaration of Helsinki, Good Clinical Practice, and relevant French regulations regarding ethics and data protection. The protocol and amendments were approved by a central independent ethics committee and written informed consent was obtained from all participants.
- Protocols to study genetics in REVERSE I was approved by the University of Toronto Research Ethics Board. Institutional research ethics board approvals were obtained by all participating centers (Ottawa Hospital Research Ethics Board).

Written informed consent to participate was obtained from all participants.

---

**Supplementary Table 1.** Imputation quality scores and minor allele frequency of ABO blood groups tagging SNPs in unprovoked-PE patients from EDITH, PADIS-PE, and REVERSE-I.

| Tagging SNPs for ABO<br>haplotypes | **EDITH**<br>N=165 |  | **PADIS-PE**<br>N=306 |  | **REVERSE-I**<br>N=115 |  |
| --- | --- | --- | --- | --- | --- | --- |
|  | R<sup>2</sup> | MAF | R<sup>2</sup> | MAF | R<sup>2</sup> | MAF |
| **rs8176719** (O1) | 0.99 | 0.39 | 0.99 | 0.49 | 0.97 | 0.45 |
| **rs41302905** (O2) | 0.99 | 0.03 | 0.99 | 0.03 | 0.83 | 0.03 |
| **rs2519093** (A1) | 0.99 | 0.24 | 0.99 | 0.33 | 0.94 | 0.29 |
| **rs1053878** (A2) | 0.99 | 0.06 | 0.99 | 0.06 | 0.98 | 0.05 |
| **rs8176743** (B) | 0.99 | 0.06 | 1.00 | 0.08 | 0.99 | 0.09 |

R<sup>2</sup>: imputation quality score
MAF: minor allele frequency

**Supplementary Table 2.** Associations of ABO haplotypes with presence^+^ of RPVO in unprovoked-PE patients from EDITH, PADIS-PE, and REVERSE-I.

| ABO<br>haplotypes | EDITH<br>N=165 |  | PADIS-PE<br>N=306 |  | REVERSE<br>N=115 |  | Meta-analysis<br>N=586 |  |
| --- | --- | --- | --- | --- | --- | --- | --- | --- |
|  | OR [95% CI] | p* | OR [95% CI] | p* | OR [95% CI] | p* | OR [95% CI] | p |
| O1 | Reference |  | Reference |  | Reference |  | Reference |  |
| O2 | 3.62 [0.52-24.93] | 0.19 | 1.05 [0.36-3.10] | 0.92 | 0.49 [0.08-2.89] | 0.43 | 1.12 [0.49-2.58] | 0.79 |
| A1 | 2.02 [1.02-4.02] | 0.04 | 0.85 [0.55-1.30] | 0.45 | 0.91 [0.42-1.98] | 0.81 | 1.06 [0.76-1.46] | 0.77 |
| A2 | 2.40 [0.71-80.08] | 0.16 | 0.58 [0.28-1.22] | 0.15 | 1.65 [0.50-5.39] | 0.41 | 0.99 [0.56-1.72] | 0.96 |
| B | 1.11 [0.32-3.82] | 0.87 | 1.63 [0.81-3.25] | 0.17 | 1.74 [0.48-6.22] | 0.39 | 1.53 [0.88-2.64] | 0.13 |

\*Associations were tested using logistic regression adjusted on age, sex and the four first principal components, under the assumption of additive allele effect. <sup>+</sup>Presence of RPVO was defined as RPVO>0.
O1, taken as the reference group, was tagged by the rs8176719-delG allele. The O2, A1, A2 and B blood groups were tagged by the rs41302905-T, rs2519093-T, rs1053878-A and rs8176743-T alleles, respectively. [OBJ]

**Supplementary Table 3.** Associations of ABO haplotypes with presence^+^ of RPVO defined by usual thresholds in unprovoked-PE patients in meta-analysis.

| ABO<br>haplotype | Meta-analysis<br>N=586 |  |  |  |  |  |
| --- | --- | --- | --- | --- | --- | --- |
|  | RPVO>0 |  | RPVO≥5 |  | RPVO≥10 |  |
|  | OR [95% CI] | p* | OR [95% CI] | p* | OR [95% CI] | p* |
| O1 | Reference |  | Reference |  | Reference |  |
| O2 | 1.12 [0.48-2.58] | 0.79 | 1.28 [0.56-2.95] | 0.56 | 0.98 [0.44-2.23] | 0.97 |
| A1 | 1.06 [0.76-1.46] | 0.77 | 1.05 [0.76-1.45] | 0.78 | 0.86 [0.61-1.21] | 0.39 |
| A2 | 0.99 [0.56-1.72] | 0.96 | 1.18 [0.69-2.04] | 0.54 | 1.11 [0.62-1.99] | 0.72 |
| B | 1.53 [0.88-2.64] | 0.13 | 1.27 [0.75-2.17] | 0.37 | 1.00 [0.57-1.78] | 0.99 |

\*P-value estimated following meta-analysis of EDITH, PADIS-PE and REVERSE-I individual association study using logistic regression adjusted on age, sex and the four first principal components, under the assumption of additive allele effect. <sup>+</sup>Presence of RPVO was defined as RPVO>0, RPVO≥5 or RPVO≥10.
O1, taken as the reference group, was tagged by the rs8176719-delG allele. The O2, A1, A2 and B blood groups were tagged by the rs41302905-T, rs2519093-T, rs1053878-A and rs8176743-T alleles, respectively. DOI

## Supplementary Analysis 1. Power analysis

A simulation study was conducted to assess the power of our study to detect a potential effect of ABO blood groups. Our simulation followed these steps.

First, we estimated the parameters required to simulate RPVO values from a Tweedie distribution, which appropriately reflects the semi-continuous nature of this phenotype (1,2). Specifically, we estimated:

- **ρ**, the index parameter defining the distribution’s shape,
- **ϕ**, the dispersion parameter, and
- **ì**, the expected individual mean based on selected covariates.

These parameters were obtained by applying a CPG model, adjusted for age, sex, and study cohort, to the combined dataset from the EDITH, PADIS-PE, and REVERSE-I studies.

Second, we simulated 1,000 datasets for various minor allele frequencies (MAFs) and Odds Ratios (OR). For each simulation settings, we created a sample of 586 individuals (a number that corresponds to the final size of our study when EDITH, PADIS-PE and REVERSE-I are combined) with genotype data generated based on the corresponding MAF, assuming Hardy-Weinberg equilibrium. RPVO values were then generated according to the CPG model defined in the first step and the fixed effect of each additional copy of the risk allele on RPVO.

Finally, a CPG model was fitted to each simulated dataset to estimate the association of alleles with RPVO. Statistical power was calculated as the proportion of simulations in which a significant association (α=0.05) was detected.

---

**Supplementary Table 4.** Power calculations to detect genetic associations with RPVO in 586 participants.

| **OR** | **MAF** |  |  |  |  |  |  |  |  |
| --- | --- | --- | --- | --- | --- | --- | --- | --- | --- |
|  | **0.03** | **0.05** | **0.10** | **0.20** | **0.25** | **0.30** | **0.35** | **0.40** | **0.45** |
| **1.10** | 0.08 | 0.10 | 0.13 | 0.18 | 0.20 | 0.23 | 0.24 | 0.27 | 0.33 |
| **1.20** | 0.13 | 0.19 | 0.33 | 0.50 | 0.60 | 0.66 | 0.71 | 0.77 | 0.80 |
| **1.30** | 0.25 | 0.36 | 0.59 | 0.84 | 0.89 | 0.93 | 0.96 | 0.97 | 0.99 |
| **1.40** | 0.37 | 0.54 | 0.81 | 0.98 | 0.99 | 0.99 | 1.00 | 1.00 | 1.00 |
| **1.50** | 0.51 | 0.73 | 0.92 | 0.99 | 1.00 | 1.00 | 1.00 | 1.00 | 1.00 |
| **1.60** | 0.66 | 0.86 | 0.98 | 1.00 | 1.00 | 1.00 | 1.00 | 1.00 | 1.00 |
| **1.70** | 0.74 | 0.93 | 1.00 | 1.00 | 1.00 | 1.00 | 1.00 | 1.00 | 1.00 |
| **1.80** | 0.83 | 0.96 | 1.00 | 1.00 | 1.00 | 1.00 | 1.00 | 1.00 | 1.00 |
| **1.90** | 0.89 | 0.98 | 1.00 | 1.00 | 1.00 | 1.00 | 1.00 | 1.00 | 1.00 |
| **2.00** | 0.92 | 0.99 | 1.00 | 1.00 | 1.00 | 1.00 | 1.00 | 1.00 | 1.00 |
| **2.30** | 0.99 | 1.00 | 1.00 | 1.00 | 1.00 | 1.00 | 1.00 | 1.00 | 1.00 |
| **2.50** | 0.99 | 1.00 | 1.00 | 1.00 | 1.00 | 1.00 | 1.00 | 1.00 | 1.00 |
| **3.00** | 1.00 | 1.00 | 1.00 | 1.00 | 1.00 | 1.00 | 1.00 | 1.00 | 1.00 |

Power was calculated over 1,000 simulated datasets.

